## Supplementary Material for "Diagnostic performance and clinical implications of rapid SARS-CoV-2 antigen testing in Mexico using real-world nationwide COVID-19 registry data"

**Supplementary Table 1**. Comparison and clinical characteristics of positive SARS-CoV-2 cases identified using rapid antigen tests and RT-PCR in Mexico.

| Parameters | RT-PCR positive SARS-CoV-2  n=527,068 | Ag-T positive SARS-CoV2-  n=1,599,793 | p-value |
| --- | --- | --- | --- |
| Age (years) | 41±16.4 | 44.5±17 | <0.001 |
| Male sex (%) | 256,251 (48.6) | 809,701 (50.6) | <0.001 |
| Diabetes (%) | 49,410 (9.4) | 233,089 (14.6) | <0.001 |
| COPD (%) | 3,120 (0.6) | 20,327 (1.3) | <0.001 |
| Asthma (%) | 9,411 (1.8) | 37,569 (2.3) | <0.001 |
| Immunosuppression (%) | 2,506 (0.5) | 14,917 (0.9) | <0.001 |
| Hypertension (%) | 66,560 (12.6) | 300,882 (18.8) | <0.001 |
| Other (%) | 7,227 (1.4) | 34,139 (2.1) | <0.001 |
| CVD (%) | 4,746 (0.9) | 27,987 (1.7) | <0.001 |
| Obesity | 56,483 (10.7) | 255,456 (16) | <0.001 |
| CKD (%) | 4,285 (0.8) | 26,952 (1.7) | <0.001 |
| Smoking (%) | 41,318 (7.8) | 113,741 (7.1) | <0.001 |
| Pneumonia(%) | 25,402 (4.8) | 272,296 (17) | <0.001 |
| Hospitalization(%) | 32,052 (6.1) | 356,856 (22.3) | <0.001 |
| ICU admission (%) | 31,143 (5.9) | 322,950 (20.2) | <0.001 |
| Death (%) | 18,275 (3.5) | 171,518 (10.7) | <0.001 |
| Time to assessment* (days) | 3 (1-5) | 4 (2-6) | <0.001 |

**SUPPLEMENTARY METHODS**

*Available variables*

Demographic and health data for all suspected COVID-19 are collected by personnel from each healthcare or testing facility and uploaded to the NESS. Variables available for all cases in the NESS include age, sex, nationality, state and municipality where the case was detected, immigration status as well as identification of individuals who speak indigenous languages from Mexico. Health information includes the status of diabetes, obesity, chronic obstructive pulmonary disease (COPD), immunosuppression, pregnancy, arterial hypertension, cardiovascular disease, chronic kidney disease (CKD), and asthma. Date of symptom onset, hospital admission, and death are available for all cases as are outpatient or hospitalized status, information regarding the diagnosis of pneumonia, ICU admission, and whether the patient required invasive ventilation. Full versions of the dataset and its descriptors are available at: <https://www.gob.mx/salud/documentos/datos-abiertos-152127>.

*Commercial qRT-PCR and Rapid Ag-T kits available in Mexico*

According to the Mexican Health Secretary, up to 6^th^ of April of 2021, there were 85 approved qRT-PCR testing kits for SARS-CoV-2 in Mexico. Each one had its own CT threshold which could be consulted within the following URL: <https://www.gob.mx/salud/documentos/listado-de-pruebas-moleculares-por-rt-pcr-monoplexado-sars-cov-2?state=published>. Additionally, up to 13^th^ of May of 2021, there were approved 9 Rapid Ag-T commercial testing kits distributed in Mexico. The full list is available in: <https://www.gob.mx/salud/documentos/listado-de-pruebas-de-antigeno-para-sars-cov-2?state=published>

**SUPPLEMENTARY FIGURES**

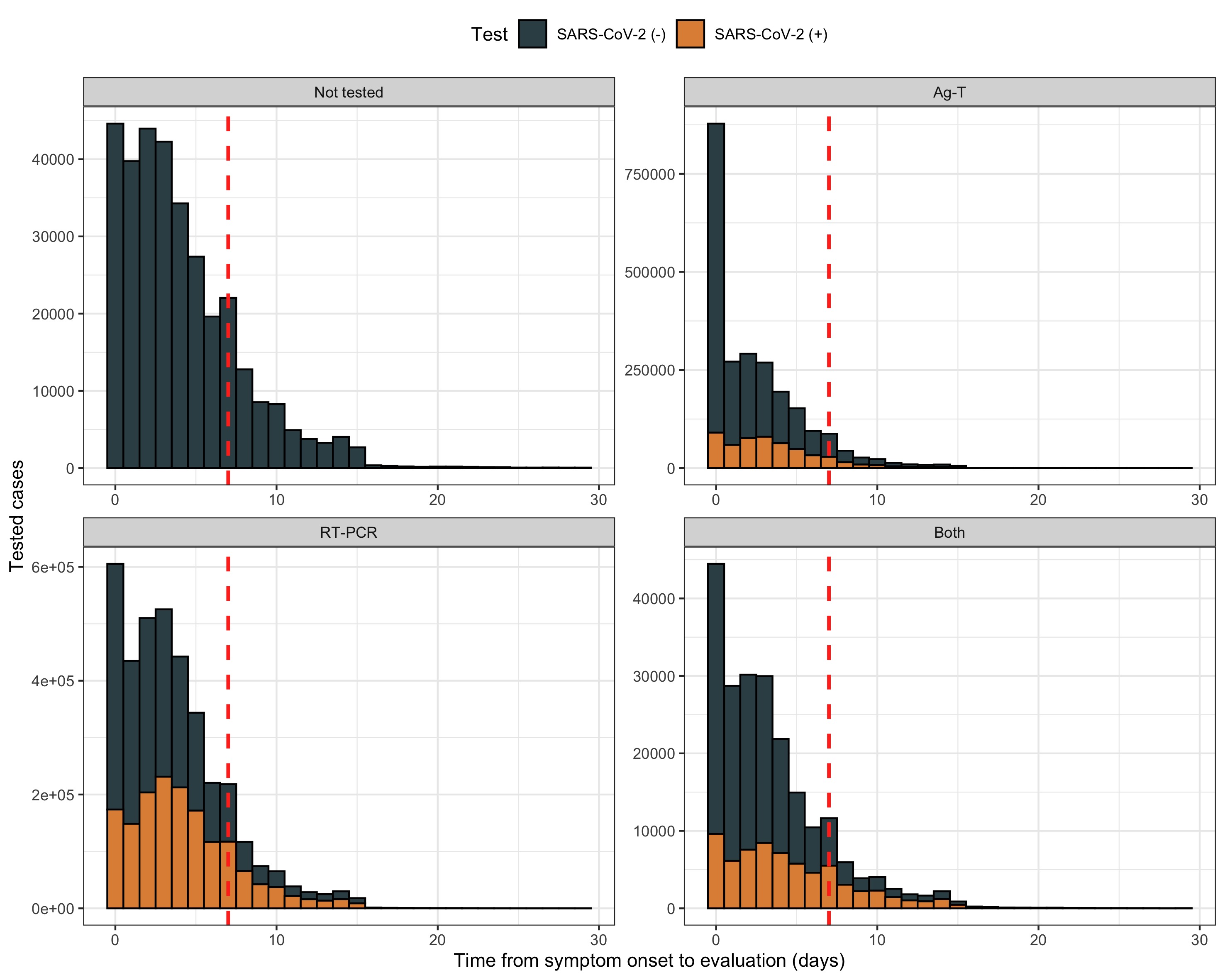

**Supplementary Figure 1**. Days from symptom onset to clinical assessment for suspected COVID-19 cases in Mexico, indicating the 7-day mark as a red dotted line.

**Index test**

n= 216,388

**Positive R-Ag-T SARS-CoV-2**

N=30,505

**Suspected SARS-CoV-2 cases**

n= 6,632,938

**Not tested (**n= 324,974)

-Contact tracing with suspected COVID-19 and no test (n= 40,893)

-Suspected COVID-19 death (n= 6,690)

-No test result available (n= 277,391)

**Negative R-Ag-T SARS-CoV-2**

N=185,883

**Eligible participant tests**

n= 6,307,964

**Reference test**

n=24,310

Target condition present

n=20,738

Target condition not present

n=3,572

**Reference test**

n=169,514

Target condition present

N=41,050

Target condition not present

n=128,464

**No index test (n=** 6,091,576)

-Only RT-PCR, n= 2,387,598

-Only rapid Ag-test, n= 3,703,978

**No reference test** (n= 6,195)

-Pending result, n= 5,839

-Inadequate sample, n= 356

**No reference test** (n= 16,369)

-Pending result, n= 12,534

-Inadequate sample, n= 3,835

**Supplementary Figure 2.** STARD diagram of case selection for evaluation of diagnostic performance of rapid antigen tests compared to RT-PCR in Mexico

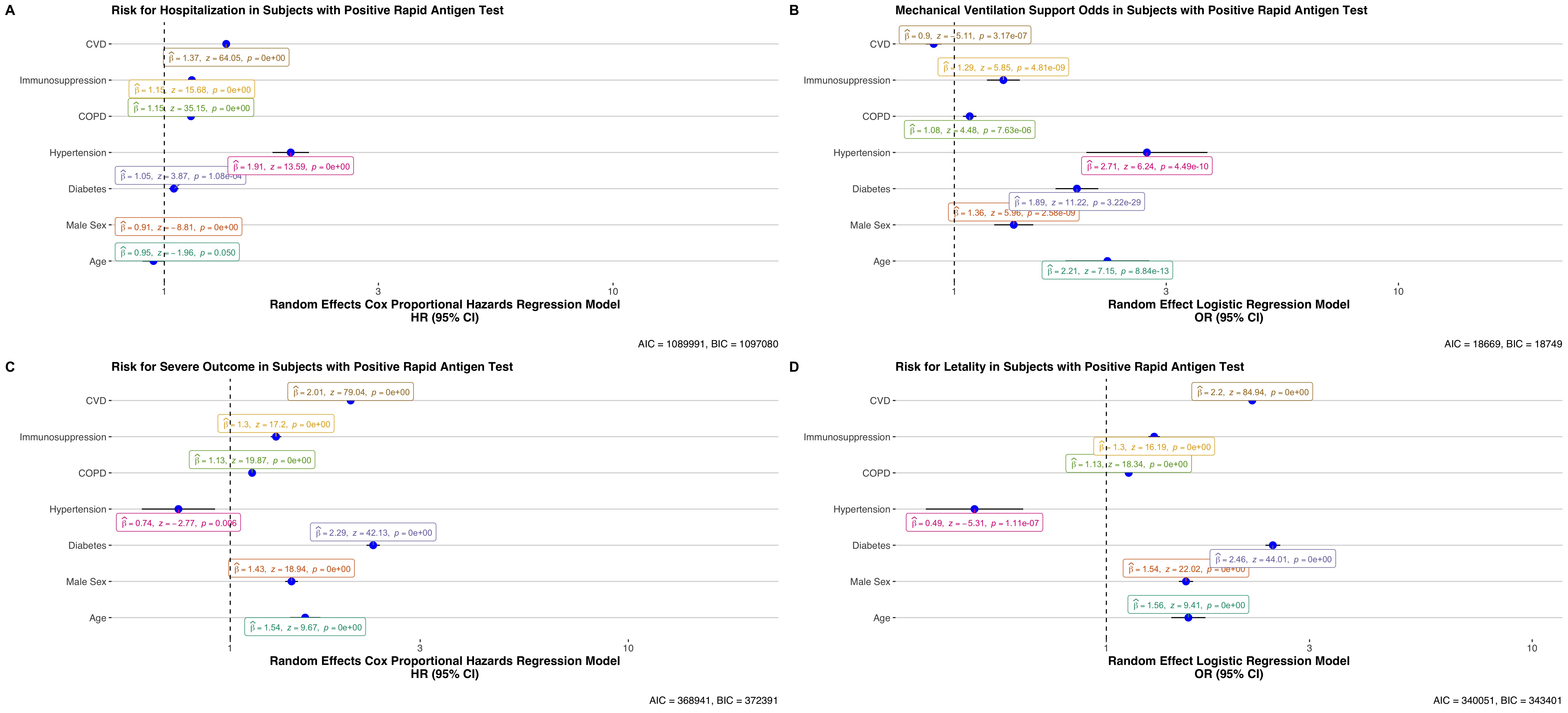

**Supplementary Figure 3.** Predictors of COVID-19 outcomes in cases identified using rapid antigen test results assessing risk for hospitalization (A), requirement for intubation (B), severe COVID-19 (C) and lethality (D) in Mexico.

**Abbreviations:** RT-PCR: Reverse transcription polymerase chain reaction; CKD, Chronic Kidney Disease; CVD: cardiovascular disease; COPD: Chronic Obstructive Pulmonary Disease; OR: Odds Ratio; HR: Hazard ratio; 95%CI: 95% Confidence interval.
